# Within- and Between-Assessor Reliability of Biarticular Muscle-Tendon Unit Lengths: Application of a Conventional Gait Model 2-based musculoskeletal model

**DOI:** 10.64898/2026.01.12.26343933

**Authors:** Cloé Dussault-Picard, Stephane Armand, Mickael Fonseca, Nathalie De Beukelaer, Morgan Sangeux, Antoine Nordez, Raphael Gross, Fabien Leboeuf

## Abstract

Cerebral palsy (CP) is characterized by neuromusculoskeletal impairments, including reduced muscle fiber lengths, which alter muscle-tendon unit (MTU) lengths and contribute to gait deviations. Estimation of MTU length reliability using musculoskeletal modeling is essential for guiding interventions such as muscle lengthening. This study aims to assess within-assessor (WA) and between-assessor (BA) reliability of MTU length estimation during gait in CP and non-impaired (NI) individuals.

38 individuals (19CP,19TD) participated in 3 3DGA sessions (2 by the same assessor). Normalized MTU length, MTU lengthening, and maximum lengthening range of the rectus femoris, semitendinosus, and gastrocnemius medialis were reported. Reliability was quantified through the standard error of measurement (SEM) and minimal detectable change (MDC).

The mean SEM (MDC) during the gait ranged from 1.0-2.1% (2.5-5.7%) for normalized MTU length, from 3.6-8.7 mm (10.1-24.1 mm) for MTU lengthening, and from 2.9-8.0 mm (8.0-22.1 mm) for MTU lengthening range in individuals with CP. In NI individuals, the mean SEM (MDC) during the cycle ranged from 0.6-1.3% (1.8-3.7%) for normalized MTU length, from 2.6-5.0 mm (7.3-13.9 mm) for MTU lengthening, and from 2.7-5.0 mm (7.5-12.4 mm) for MTU lengthening range.

Results suggest reliable estimations by the same assessor, supporting therapeutic decision-making and patient progress monitoring.

## 1. Introduction

Cerebral palsy (CP) encompasses neuromotor disorders affecting 1.6 birth per 1000 children in developed countries ^1^. CP is associated with progressive neuromusculoskeletal impairments, such as bony torsions (i.e., altered muscle moment arms), contractures (i.e., reduced muscle fiber length) ^2^, lack of selectivity ^3^ and spasticity ^4^. These neuromusculoskeletal impairments contribute to walking deviations ^5^, limited participation ^6^ and reduced functional independence ^7,8^ compared to non-impaired (NI) peers.

Three-dimensional gait analysis (3DGA) plays a crucial role in identifying the neuromusculoskeletal impairments that affect walking in individuals with CP ^9^. However, it does not provide information about changes in muscle-tendon unit (MTU) length, that could be used to analyse MTU kinematics and contraction modalities. While joint kinematics measured during movements can implicitly inform on mono-articular MTU length by reflecting changes in joint angles that are directly affected by these muscles, estimation of bi-articular MTU length requires to use musculoskeletal models. Understanding the behavior of bi-articular muscles is a key factor in clinical decision-making to determine muscle lengthening interventions ^10–13^. This measurement is accessible through the use of musculoskeletal software ^14,15^ or libraries ^16^.

Kainz et al. ^17^ advocated for the integration of musculoskeletal modeling into routine practice of clinical gait analysis. They proposed a framework based on an OpenSim model, driven by joint angles derived from the Conventional Gait Model (CGM). They further emphasized the importance of adhering to a consistent workflow when implementing musculoskeletal models. However, the estimation of MTU length in a clinical setting — for instance in pre- and post-muscle lengthening evaluations — can be confounded by intrinsic (e.g., the individual’s ability to walk consistently in the same manner) and extrinsic variability (e.g., marker placement errors, measurement technique).

To our knowledge, no studies have reported the reliability of MTU length estimation in musculoskeletal models within a clinical context. Our study aimed to address this gap by examining the reliability of MTU length estimations. Drawing on the recommendations of McGinley et al. ^18^, who advised the use of unit-based reliability metrics such as the standard error of measurement (SEM), we aimed to report both between-assessor (BA) and within-assessor (WA) SEM and minimal detectable change (MDC) values for individuals with CP and their NI peers.

## 2. Methods

### 2.1. Participants

This study retrospectively assessed gait gata of participants aged between 6 and 43 years old recruited at the Geneva University Hospitals. The exclusion criteria for both groups were known pregnancy and allergy to adhesive tape. Individuals were included in the CP group if they had a confirmed diagnosis of CP, the ability to walk 10 meters without assistive aid, and the capacity to understand and follow verbal instructions. The NI individuals were included if they had no pathological condition affecting typical motor ability. The research protocol was approved by the “Commission Cantonale d’Éthique de la Recherche Genève” (CCER-2020-00358) and all the participants or their legal tutors (for non-adult participants) provided written informed consent.

### 2.2. Experimentation

Participants visited the laboratory on two separate occasions (S1 and S2), spaced 10 days apart. On the first visit (S1), a 3DGA session was conducted by one assessor (A1) (S1-A1). On the second visit (S2), two consecutive 3DGA sessions were conducted by A1 and A2 (S2-A1 and S2-A2), respectively. Each 3DGA session included a minimum of 10 barefoot walking trials at a self-selected comfortable speed. Each assessor was responsible for placing the markers for the 3DGA they were conducting. Both assessors had extensive experience in identifying anatomical landmarks. Reflective markers (14 mm) were placed following the CGM (version 2.3) ^19^, and palpation followed the guidelines described by Van Sint Jan (2007) ^20^. The marker trajectories were recorded at 100 Hz by a 12-camera motion capture system (Oqus7+, Qualisys, Göteborg, Sweden).

### 2.3. Data processing

Initial data processing (i.e., marker labelling and gap filling, foot strike and foot-off detection) was conducted in QTM (Qualisys, Göteborg Sweden). The generic Gait2392 OpenSim model was scaled using the marker positions of a standing static trial ^21^. After this calibration step, a scaled musculoskeletal model was generated. The scaling parameters (geometric bones scales and marker placer weights) are presented in **supplementary file**. Then, kinematic fitting and joint kinematics were conducted using the CGM2.3 version of the open source library pyCGM2 ^19^. The scaled OpenSim model was joint driven to estimates MTU lengths of 92 muscles. Joint kinematics and MTU lengths were time-normalized to 101 data points according to foot contacts. The MTU lengths of 8 biarticular muscles of the lower limbs were retained for further analysis (semimembranosus, semitendinosus, biceps femoris, rectus femoris, gastrocnemius medialis, gastrocnemius lateralis, sartorius, and gracilis). All the available curves were averaged for each leg and muscle. Only the impaired legs (n = 29) were kept for the CP individuals (unilateral: n = 9; bilateral: n = 10). To avoid functional redundancy, only the results of the rectus femoris, semitendinosus, and gastrocnemius medialis were reported. The results of the other biarticular muscles are provided in supplementary file.

### 2.4. MTU length normalization

At each frame (*i*) of the gait cycle, the normalized MTU length (𝐿𝑒𝑛𝑔𝑡ℎ(𝑖)) was calculated as the MTU length (*Length(i)*) normalized to the reference length in the virtual standing position, i.e. with all joint coordinates set to 0° (*Anatomical MTU Length*) ^22^:

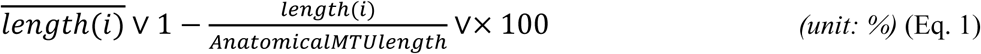

### 2.5. Muscle-tendon lengthening

The MTU lengthening at each instant *i* ((*Lengthening(i))* was calculated as the length difference between the MTU length (*Length(i)*) subtracted by the *Anatomical MTU Length:*

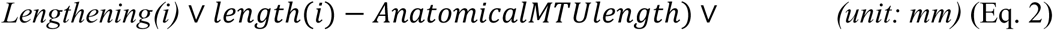

We extracted from eq.2, the maximum lengthening range throughout the gait cycle as:

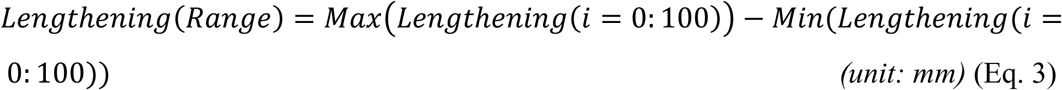

### 2.6. Statistical analysis

The standard error of measurement (SEM) was calculated; on a frame-by-frame basis for the normalized MTU length (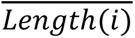 and the MTU lengthening *i* (*Lengthening(i)),* for each biarticular muscle, using the residual standard deviation (σ*r*) from an analysis of variance (ANOVA) model:

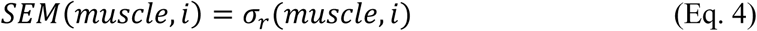

The minimal detectable change (MDC) was calculated for each muscle as:

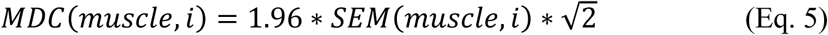

The *summary* 𝑆𝐸𝑀and 𝑀𝐷𝐶 were calculating by averaging the parameters across the gait cycle, to provide one value for each biarticular muscle.

Additionally, the 𝑆𝐸𝑀 and 𝑀𝐷𝐶 for the lengthening range (*Lengthening(Range))*, i.e., a discrete value calculated for each gait cycle, were estimated from a similar processing based on a variance analysis.

All statistical analysis steps were run in RStudio (v.2024.04.2, PBC, Boston, MA).

## 3. Results

### 3.1. Participants

A total of 19 individuals with CP (Gross Motor Function Classification System (GMFCS) I: n = 15, GMFCS II: n = 3, GMFCS III: n = 1) and 19 NI individuals were recruited. Anthropometrics and characteristics of each group are reported in **Table 1**.

**Table 1.** Participants’ characteristics.

|  | **CP** | **NI** |
| --- | --- | --- |
| **n** | 19 | 19 |
| **Age (years)** | 18.4 (7.3) | 18.3 (11.2) |
| **Sex n (F/M)** | 5F / 14M | 9F / 10M |
| **BMI (kg.m<sup>-2</sup>)** | 21.0 (4.21) | 19.3 (3.7) |
| **Height (m)** | 1.61 (0.16) | 1.52 (0.18) |
| **Weight (kg)** | 56.1 (17.9) | 46.9 (18.1) |
| **GMFCS level** | I: n = 15 |  |
|  | II: n = 3 | N/A |
|  | III: n = 1 |  |
| **Involvement** | Unilateral: n = 9 | N/A |
|  | Bilateral: n = 10 |  |

Mean (standard deviation) of participant anthropometrics and characteristics for the cerebral palsy (CP) and the non-impaired (NI) groups. Sample size (n), side of the muscle disorder involvement; unilateral (U) or bilateral (B), or not applicable (N/A), sex; female (F) and male (M), and Gross Motor Function Classification System (GMFCS) level are also reported.

### 3.2. Normalized muscle-tendon unit length

In the CP group, the highest values of normalized MTU length during the gait cycle were 8.2 ± 2.5% for the rectus femoris, 9.2 ± 1.9% for the semitendinosus, and 6.7 ± 2.3% for the gastrocnemius medialis (**Figure 1a, first row**). In the NI group, the highest values were 8.9 ± 1.6% for the rectus femoris, 10.0 ± 1.5% for the semitendinosus, and 8.9 ± 1.9% for the gastrocnemius medialis (**Figure 2a, first row**).

**Figure 1.**
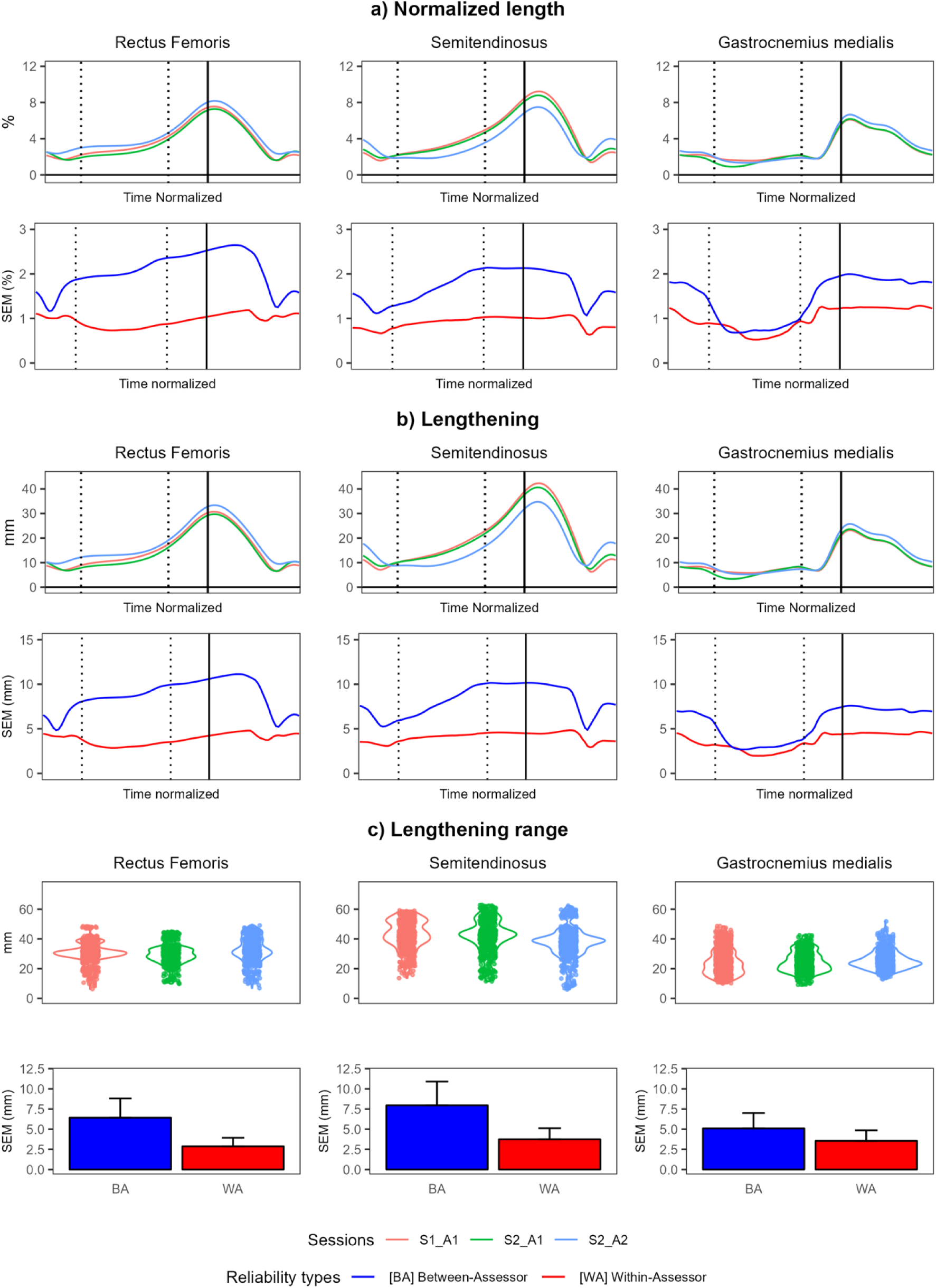
Time-normalized results of the cerebral palsy group for the three sessions (session 1 – assessor 1 (S1_A1), session 2 – assessor 1 (S2-A1, and session 2 – assessor 2 (S2_A2)) and associated within-(WA) and between-assessor (BA) standard error of measurement (SEM) for the rectus femoris (left column), the semitendinosus (center column) and the gastrocnemius medialis (right column). Results are presented for MTU a) normalized length, b) lengthening, and c) lengthening range. The dotted vertical lines indicate the beginning and end of the single stance phase, while the solid vertical line represents toe-off, marking the transition to the swing phase.

**Figure 2.**
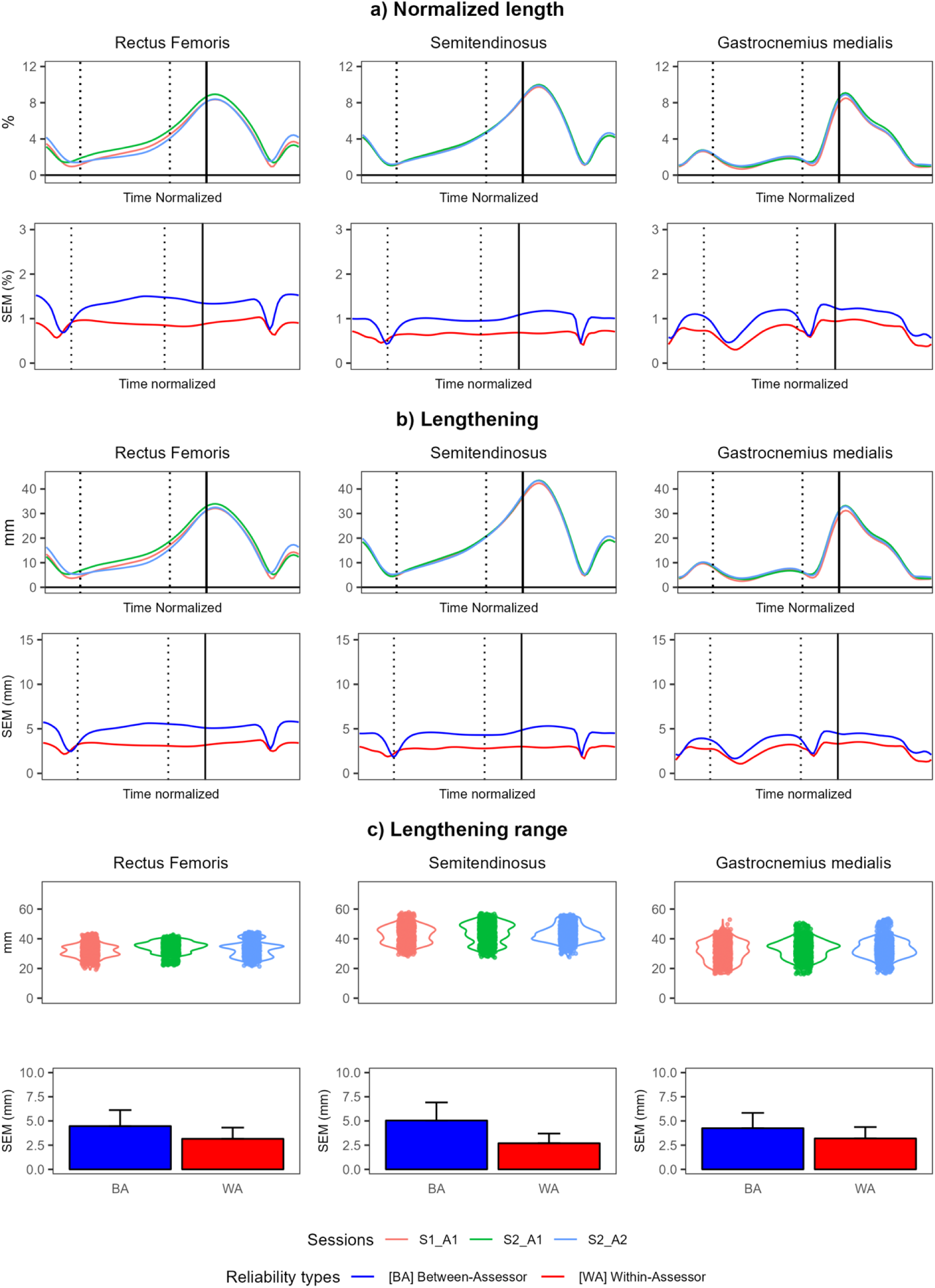
Time-normalized results of the non-impaired group for the three sessions (session 1 – assessor 1 (S1_A1), session 2 – assessor 1 (S2-A1, and session 2 – assessor 2 (S2_A2)) and associated within-(WA) and between-assessor (BA) standard error of measurement (SEM) for the rectus femoris (left column), the semitendinosus (center column) and the gastrocnemius medialis (right column). Results are presented for MTU a) normalized length, b) lengthening, and c) lengthening range. The dotted vertical lines indicate the beginning and end of the single stance phase, while the solid vertical line represents toe-off, marking the transition to the swing phase.

Concerning BA reliability, the rectus femoris presented the highest mean SEM values for normalized length in both groups (CP-SEM 2.1%, CP-MDC: 5.7%; NI-SEM: 1.3%, NI-MDC: 3.7%), with higher values in the CP group (Table 2). The SEM reached its peak during mid-swing (2.6%) in the CP group (**Figure 1a, second row)**, and at the end of swing phase (1.5%) in the NI group (**Figure 2a, second row**). The group mean and standard deviation of maximal values reached during the gait cycle are presented in **supplementary material**.

**Table 2.** Standard error of measurement and minimal detectable change results.

|  |  | Normalized length |  |  |  | Lengthening |  |  |  | Lengthening range |  |  |  |
| --- | --- | --- | --- | --- | --- | --- | --- | --- | --- | --- | --- | --- | --- |
|  |  | CP |  | NI |  | CP |  | NI |  | CP |  | NI |  |
| Muscle |  | SEM (%) | MDC (%) | SEM (%) | MDC (%) | SEM (mm) | MDC (mm) | SEM (mm) | MDC (mm) | SEM (mm) | MDC (mm) | SEM (mm) | MDC (mm) |
| Between assessor | Semitendinosus | 1.7 | 4.8 | 1.0 | 2.7 | 8.2 | 22.7 | 4.4 | 12.2 | 8.0 | 22.1 | 5.0 | 6.9 |
|  | Rectus femoris | 2.1 | 5.7 | 1.3 | 3.7 | 8.7 | 24.1 | 5.0 | 13.9 | 6.4 | 17.8 | 4.5 | 12.4 |
|  | Gastrocnemius medialis | 1.5 | 4.0 | 1.0 | 2.7 | 5.6 | 15.6 | 3.5 | 9.8 | 5.1 | 14.2 | 4.3 | 11.8 |
| Within assessor | Semitendinosus | 1.0 | 2.5 | 0.6 | 1.8 | 4.1 | 11.3 | 2.8 | 7.7 | 3.7 | 10.4 | 2.7 | 7.5 |
|  | Rectus femoris | 1.0 | 2.7 | 0.9 | 2.4 | 3.8 | 10.7 | 3.2 | 8.9 | 2.9 | 8.0 | 3.2 | 8.7 |
|  | Gastrocnemius medialis | 1.0 | 2.8 | 0.7 | 2.0 | 3.6 | 10.1 | 2.6 | 7.3 | 3.6 | 9.8 | 3.2 | 8.8 |

Mean within and between-assessor standard error of measurement (SEM) and minimal detectable change (MDC) across the gait cycle for each biarticular muscle in the cerebral palsy (CP) and the non-impaired (NI) groups.

The WA-SEM values were lower than BA-SEM for all muscles in both groups (**Figure 1a, 2a, second row**), and more similar across muscles (rectus femoris: CP-SEM: 1.0%, CP-MDC: 2.7%; NI-SEM: 0.9%, NI-MDC: 2.4%; semitendinosus: CP-SEM: 0.9%, CP-MDC: 2.5%; NI-SEM: 0.6%, NI-MDC: 1.8%; gastrocnemius medialis: CP-SEM 1.0%, CP-MDC: 2.8%; NI-SEM: 0.7%, NI-MDC: 2.0%) (**Table 2**). Normalized MTU length of all other biarticular muscles (biceps femoris, semimembranosus, gastrocnemius lateralis, gracilis and sartorius) are presented in **supplementary material**.

### 3.3. Muscle-tendon unit lengthening

The semitendinosus exhibited the highest MTU lengthening among the three muscles in both groups (CP: 42.3 ± 9.6 mm; NI: 43.5 ± 7.0 mm) (**Figure 1b, 2b, first row)**.

Concerning the BA reliability, the rectus femoris showed the largest mean SEM during the gait cycle for lengthening in both groups (CP-SEM: 8.7 mm, CP-MDC: 24.1 mm; NI-SEM: 5.0 mm, NI-MDC: 13.9 mm), with higher values in the CP group (**Table 2**). The SEM reached its peak during mid-swing (11.1 mm) in the CP group (**Figure 1b, second row)**, and at the end of swing phase (5.8 mm) in the NI group (**Figure 2b, second row**).

The WA-SEM values were lower than BA-SEM for all muscles in both groups (**Figure 1b, 2b, second row**), and more similar across muscles (rectus femoris: CP-SEM 3.8 mm, CP-MDC: 10.7 mm; NI-SEM: 3.2 mm, NI-MDC: 8.9 mm; semitendinosus: CP-SEM 4.1 mm, CP-MDC: 11.3 mm; NI-SEM: 2.8 mm, NI-MDC: 7.7 mm; gastrocnemius medialis: CP-SEM: 3.6 mm, CP-MDC: 10.7 mm; NI-SEM: 2.6 mm, NI-MDC: 7.3 mm) (**Table 2**).

### 3.4. Muscle-tendon unit lengthening range

The semitendinosus exhibited the highest lengthening range among the three muscles in both groups (CP: 42.8 ± 9.6 mm; NI: 43.5 ± 5.6 mm) (**Figure 1c, 2c, first row)**.

The mean BA-SEM during the gait cycle remains similar across muscles (rectus femoris: CP-SEM: 6.4 mm, CP-MDC: 17.8 mm; NI-SEM: 4.5 mm, NI-MDC: 12.4 mm; semitendinosus: CP-SEM: 8.0 mm, CP-MDC: 22.1 mm; NI-SEM: 5.0 mm, NI-MDC: 6.9 mm; gastrocnemius medialis: CP-SEM: 5.1 mm, CP-MDC: 14.2 mm; NI-SEM: 4.3 mm, NI-MDC: 11.8 mm) (**Table 2**), with higher values in the CP groups (**Figure 1c, 2c, second row**).

Again, the WA-SEM values were lower than BA-SEM (**Figure 1c, 2c, second row**) for all muscles in both groups (rectus femoris: CP-SEM: 2.9 mm, CP-MDC: 8.0 mm; NI-SEM: 3.2 mm, NI-MDC: 8.7 mm; semitendinosus: CP-SEM: 3.7 mm, CP-MDC: 10.4 mm; NI-SEM: 2.7 mm, NI-MDC: 7.5 mm; gastrocnemius medialis: CP-SEM: 3.6 mm, CP-MDC: 9.8 mm; NI-SEM: 3.2 mm, NI-MDC: 8.8 mm) (**Table 2**).

## 4. Discussion

Overall, the mean SEM during the gait cycle ranged from approximately 1.0 to 2.1% for normalized MTU length and 3.6 to 8.7 mm for MTU lengthening in individuals with CP. In general, higher SEM values were noted in the CP compared to the NI group, likely reflecting the greater variability in movement patterns known in this population ^23^. Our SEM findings align with those of Hanssen et al. ^24^, who measured MTU lengths (normalized to segment length) in a static position using 3D ultrasound. They reported SEM-WA values for the rectus femoris of 1.5% and 2.0%, and for the gastrocnemius medialis of 1.2% and 0.9% in the CP and NI populations, respectively ^24^. This concordance suggests that MTU length estimations derived from musculoskeletal modeling could offer reliability comparable to 3D ultrasound for tracking relative changes across repeated measurements. Future studies would compare MTU length estimated using musculoskeletal models and imaging and assess whether imaging would help to personalize models and improve their validity. This step would be particularly relevant for patients with bone deformations. However, SEM-WA findings of MTU lengthening were slightly higher than ultrasound reliability investigated by Guenanten et al. ^25^, who reported SEM (MDC) values of 2 mm (6 mm) for the hamstrings, and 1 mm (3 mm) for other muscles in adults. This discrepancy may be attributed to differences in methodology, as musculoskeletal modeling involves multiple sources of variability (e.g., marker placement, model scaling, and joint kinematics estimation) and is conducted during dynamic gait cycles, whereas ultrasound measurements are typically acquired in a static position.

Time-series curves such as normalized MTU lengths and MTU lengthening offer a dynamic perspective that complements static measures (e.g., passive range of motion) and can enrich clinical interpretation. By comparing these gait-derived MTU lengths to values obtained from analytically driven joint angles in a musculoskeletal model, clinicians can, within known bounds of measurement variability, better determine whether a functional reserve remains accessible to the patient. This approach offers a new layer of clinical insight: not only confirming the presence of contractures, but also assessing the mobilization potential of the remaining muscle length, as recently proposed by De Beukelaer et al. ^26^. This clinical insight may prove especially valuable, as Wren et al. ^10^ and Rajagopal et al. ^11^ emphasized that surgical lengthening yields better outcomes when targeting muscles that are reliably identified as “short”.

To justify surgical intervention, it is crucial to rely on repeatable MTU length measurements that accurately reflect the lack of lengthening during gait; thus, quantifying their repeatability is a key prerequisite for informed clinical decision-making. The mean SEM-WA of MTU lengthening during the gait cycle ranged between 3.6 and 4.1 mm, with mean MDCs of 11.3 mm for the semitendinosus, 10.7 mm for the rectus femoris, and 10.1 mm for the gastrocnemius medialis in CP individuals. In the context of clinical intervention, the obtained MDC remained below the range of cadaveric musculotendinous lengthening observed for the gastrocnemius, which varies between 15 and 26 mm depending on the level of recession ^27,28^, thus confirming the ability of MTU estimations through musculoskeletal modeling to detect clinically relevant post-lengthening changes.

A mean MDCs-WA of approximately 10 mm was reported. Subsequent analysis based on the mean joint- driven simulation-based estimates of MTU lengthening in response to imposed ranges of motion for the CP group have been conducted (see in **supplementary file)**. Results showed that this 10-mm lengthening is achieved with 11.7° of hip flexion for the semitendinosus, 14.6° of knee flexion for the rectus femoris, and 13.3° of ankle dorsiflexion for the gastrocnemius medialis. This lengthening-angle correspondence may provide clinically relevant insights for surgical planning.

Overall, SEM values were consistently higher in the CP compared to the NI group, and greater for BA than WA estimations. This pattern aligns with previous reliability studies on gait kinematics and kinetics such as Kainz et al., which reported lower standard deviations in NI individuals relative to those with CP ^29^. Similar findings were also reported by McGinley et al. ^18^ and Manca et al. ^30^, reinforcing the idea that neuromuscular variability and marker placement inconsistency contribute significantly to measurement error in clinical gait analysis.

The primary source of error is well documented and stems from the assessor’s ability to reliably reposition markers on anatomical landmarks across sessions ^31^. This source of variability can be further amplified by model scaling procedures, as emphasized by Koller et al. ^32^, especially when marker placement deviates from anatomical references. To mitigate this, it is strongly recommended that a single, trained examiner—experienced in anatomical palpation—perform all assessments. Results of this study shown that involving two assessors substantially increase SEM and MDC values, which in turn may mask the effects of interventions, particularly those involving muscle-tendon lengthening.

Several methods for normalizing MTU length during gait are commonly used in the literature, each with distinct implications for the interpretation of results. One such approach, as applied in this study, is normalizing to the length of the muscle in the anatomical position ^22^. While this method is straightforward, it may underestimate the MTU length of long muscles or those already stretched in the standing position. This method was chosen because the study aimed to provide easily interpretable SEM data and given that it is the most commonly used approach in studies involving individuals with CP ^13,17,33–38^. In contrast, other normalization methods, such as normalizing to maximal muscle lengthening (i.e., the maximal length achieved during walking) ^13,34^, may provide an alternative reflection of the muscle’s potential elongation. However, this method could overestimate the length of muscles that are not fully stretched during walking.

All SEM and MDC values reported in this study should be interpreted cautiously when applied to other workflows ^17^. In the current study, the CGM2.3 model was employed, which uses kinematic fitting with ball-and-socket joint representations, combined with OpenSim Gait2392 muscle geometry. The use of alternative muscle geometries or incorporating additional joint degrees of freedom, known to increase the joint angle variability ^39^ could potentially affect our results. Moreover, this study has several limitations. First, it evaluated only the repeatability of musculoskeletal model estimations and did not address its trueness. Assessing trueness would require a reference standard, such as medical imaging to faithfully reproduce muscle pathways ^25^. Second, our CP population included mostly individuals with high functional levels (GMFCS I: n = 15, GMFCS II: n = 3, GMFCS III: n = 1). Consequently, the findings cannot be generalized to subjects with low gross motor function (e.g., GMFCS level III). Third, this study reported SEM and MDC values separately for the CP and NI groups; however, no statistical comparison between groups was conducted. Such comparisons were not feasible, as SEM is calculated at the group level and does not yield individual values from which a mean and standard deviation could be derived. Since the objective was to provide reference values for each group rather than to compare them and given that only two within-assessor and two between-assessor evaluations were available, the use of SEM was considered the most appropriate method for assessing measurement reliability. Finally, musculoskeletal modeling does not allow to determine whether the changes in length occurs within the muscle and/or the tendon, requiring ultrasound imaging to accurately distinguish the contributions of each component ^40^.

## 5. Conclusion

In conclusion, this study helps bridge the gap between musculoskeletal modeling and clinical practice. We reported an average SEM during the cycle ranging from 1.0 to 2.1% for normalized MTU length and 3.6 to 8.7 mm for MTU lengthening in individuals with CP, which can support therapeutic decision-making and patient progress monitoring. Further studies would compare musculoskeletal modelling with imaging to better appraise the validity of the models, and potentially better individualize these models.

## Data Availability

All data produced in the present study are available upon reasonable request to the authors

## Author contributions

- Cloé Dussault-Picard (C.D.-P.): Methodology; Formal analysis; Writing – original draft; Writing – review & editing.
- Stéphane Armand (S.A.): Investigation; Writing – review & editing.
- Mickaël Fonseca (M.F.): Investigation; Writing – review & editing.
- Nathalie De Beukelaer (N.D.B.): Writing – review & editing.
- Morgan Sangeux (M.S.): Writing – review & editing.
- Antoine Nordez (A.N.): Writing – review & editing.
- Raphaël Gross (R.G.): Writing – review & editing.
- Fabien Leboeuf (F.L.): Conceptualization; Methodology; Software; Data curation; Formal analysis; Writing – review & editing; Supervision.

## Data availability statement

The datasets analysed during the current study are not publicly available due to institutional policy but are available from the corresponding author on reasonable request.

